# A Longitudinal Assessment of Adiposity Change on the Incidence of Different Cardiometabolic Diseases Among Adult Qataris

**DOI:** 10.1101/2025.08.01.25332691

**Authors:** Rawan Ajeen, Karam I. Turk-Adawi, Alice S. Ammerman, John A. Batsis, Shu Wen Ng, Linda S. Adair

## Abstract

Anthropometric and body composition changes may contribute differently to cardiometabolic disease risk depending on fat distribution and sex. Few studies have examined these longitudinal changes in Middle Eastern populations. To evaluate how five-year changes in weight, visceral adipose tissue (VAT), waist circumference (WC), and waist-to-hip ratio (WHR) relate to incident elevated blood pressure (EBP), diabetes, and dyslipidemia in Qatari adults, and whether baseline adiposity modifies these associations. Data were drawn from 1,765 Qatari adults (755 females, 1,010 males) in the Qatar Biobank cohort with repeated measures over five years. Sex-stratified logistic regression models were used to assess how changes in adiposity measures predicted incident disease. Interaction terms tested effect modification by baseline adiposity. Participants had a mean baseline age of 40 years. Dyslipidemia was highly prevalent at baseline (68% in males, 63% in females), while incidence of EBP and diabetes was relatively low. VAT gain was associated with higher odds of incident EBP in both sexes (OR: 1.53, 95% CI: 1.19–1.97 for males; OR: 1.85, 95% CI: 1.11–3.11 for females), and with diabetes in males (OR: 1.61, 95% CI: 1.14–2.25). WC change in females was associated with dyslipidemia (OR: 1.06, 95% CI: 1.02–1.10), while weight gain was inversely associated with dyslipidemia among females with obesity (OR: 0.89, 95% CI: 0.81–0.98). WHR in males was associated with both EBP and dyslipidemia, suggesting it may capture VAT-related risk. Some associations varied by baseline adiposity, with stronger effects among those with lower baseline VAT or WC. The relationship between adiposity changes and cardiometabolic risk varies by disease, sex, and baseline adiposity. WHR and WC may serve as useful proxies for VAT in risk assessment, especially in resource-limited settings.

## Introduction

Cardiovascular disease (CVD) remains the leading cause of mortality worldwide, with rates expected to rise further due to aging populations and increasing sedentary lifestyles(1). This global trend is particularly pronounced in the Middle East, where rapid urbanization has spurred a nutrition transition marked by greater dietary intake high in trans-fat and sugar and decreased physical activity(2,3). In Qatar, one of the fastest-developing nations in the region, CVD accounts for nearly 30% of all mortality, and obesity rates continue to climb, with projections suggesting that over half of adults will be living with obesity by 2035(4,5). These shifts highlight the urgent need for population-specific strategies to prevent and manage chronic disease.

Although obesity is a well-established risk factor for CVD, the relationship between adiposity and cardiometabolic disease risk is not solely dependent on total body fat. Rather, fat distribution, particularly the accumulation of visceral adipose tissue (VAT), plays a crucial role due to its pro-inflammatory and insulin-resistant properties, and secretion of cytokines and adipokines that contribute to systemic inflammation, dyslipidemia, and hypertension(6–8). These effects are especially pronounced with aging, when fat is typically redistributed toward central regions and lean muscle mass declines(9–11). However, changes in VAT are difficult to quantify without imaging techniques. Previous research suggests that abdominal anthropometric measures like waist circumference (WC) and waist-to-hip ratio (WHR) may provide accessible, cost-effective proxies for VAT(12–14).

The biological pathways linking adiposity to cardiometabolic outcomes differ by disease. Composite scores such as the Framingham Risk Score (FRS) or definitions such as Metabolic Syndrome (MetS), though commonly used, may obscure these distinctions(15). Hypertension is often driven by endothelial dysfunction while diabetes relates more to insulin resistance and ectopic fat deposition along with decreased lean mass(6–8,16,17). Dyslipidemia involves disruptions in hepatic lipid metabolism(6,18). Given these divergent mechanisms and differing clinical treatments, analyzing each outcome separately provides clearer insights into how adiposity contributes to disease risk and supports more tailored prevention strategies.

Fat distribution and its health implications differ by sex. Males typically accumulate more VAT earlier in adulthood, increasing their risk for VAT-related disease risk earlier.(19,20) Females shift from gynoid to android fat distribution during menopause, leading to greater VAT accumulation and risk with age(9,21,22). These sex-specific patterns complicate the use of general obesity metrics like BMI, which may fail to account for sex-specific nuances in fat distribution and lean mass. Assessing anthropometrics most correlated with VAT for each sex, such as WHR in males and WC in females, may better capture disease risk in targeted populations. Importantly, the relationship between changes in fat distribution and disease risk may depend on an individual’s existing adiposity(23,24). Individuals with higher initial VAT or abdominal obesity may already have heightened metabolic disturbances, such that additional fat would have a different effect on their risk compared to individuals with lower initial baseline VAT.

Over a five-year period, we investigated how changes in weight, VAT, WHR and WC were related to incident risk defined using standard cut-points for elevated blood pressure (EBP), diabetes, and dyslipidemia in a Qatari adult sample. We hypothesized that the associations between changes in adiposity and disease risk would differ across specific cardiometabolic outcomes, EBP, diabetes, and dyslipidemia, reflecting distinct biological pathways. We also anticipated that these associations would vary by sex and depend on individuals’ baseline adiposity levels. To explore potential effect modification, we formally tested interactions between baseline adiposity and changes in body composition. Understanding these relationships has important implications for public health efforts and interventions aimed at reducing the CVD burden in the region, particularly as aging and lifestyle factors continue to shift population health trends. Data on the longitudinal impacts of body composition changes on incident disease risk are limited in Qatar, highlighting the need for this study.

## Methods

### Study Sample

This study utilized longitudinal data from the Qatar BioBank (QBB) cohort study, a large-scale, prospective, population-based study focusing on Qatari national and long-term (≥ 15 years) Qatari residents. Detailed descriptions of QBB’s sampling methodology, as well as inclusion and exclusion criteria, are available in a prior publication(25). The current sampling methods of the QBB do not allow for a representative sample of the entire Qatari population(26).

Following data use agreements between QBB and Qatar University, a randomly selected subset of QBB participants (n = 6,000) was selected. Preliminary analyses using the full cohort (n = 17,065) suggest this sample is representative in age distribution, sex ratio, and health outcomes. Our study excluded pregnant women and non-Qatari nationals. For longitudinal analyses, the analytical sample included 755 females and 1,010 males who had completed a follow-up visit at least five years after their baseline assessment. Throughout this paper, we refer to Qatari nationals as Qatari’s.

The QBB study protocol received ethics approval from the Hamad Medical Corporation Ethics Committee, with all participants providing informed consent for their de-identified data to be used in research. Additional ethical approval for this study was obtained from the institutional review board of QBB (QF-QBB-RES-ACC-00217) and an exemption from the University of North Carolina at Chapel Hill IRB (#23-2942). This study was carried out according to the Declaration of Helsinki.

### Variables

#### Participant Characteristics

Sociodemographic data gathered through a self-administered questionnaire included age and sex. Health-related information including smoking status (current smoker or not), physical activity, and medication use were collected through face-to-face interviews conducted by a trained nurse.

#### Exposure Variables: Anthropometrics and Body Composition

The primary exposures were five-year changes in weight, VAT, WC, and WHR. These measures were chosen based on prior evidence of their associations with cardiometabolic disease risk(6,12,14,27–31). All exposure variables were assessed once by trained nurses using standardized measurement protocols. While wearing light clothes and no shoes(25), body weight (kg) and height (cm) were measured using a calibrated scale and a wall-mounted Seca 217 stadiometer (Seca GmbH, Hamburg, Germany)(32). WC was measured in centimeters halfway between the lower ribs and the iliac crest, while HC was measured in centimeters at the largest circumference around the buttocks^57^. BMI (kg/m^2^)(33), WHR (WC/HC)(12), and WHtR (WC/height) were calculated(34). BMI categories were defined using international standards as underweight (<18.5 kg/m^2^), normal (18.5 – 24.9 kg/m^2^), overweight (OW) 25–29.9 kg/ m^2^, and obesity as ≥30.0 kg/ m^2(^(33)^)^ . In analyses, the underweight and normal weight were grouped together due to the small sample size of the underweight category.

Body composition, including total body fat percentage (%BF) was measured by a full body General Electric Lunar dual energy X-ray absorptiometry (DXA) scan (GE Healthcare, Madison, WI)(35). Participants were instructed to lay flat and still on the scanning table while wearing a light gown and no jewelry(25). CoreScan software was used to estimate VAT from DXA(36). A previous study conducted on a similar population found that VAT estimates from CoreScan were comparable to that obtained from the gold standard MRI or CT scan(37–41). VAT percentages were calculated as relative to total %BF and as absolute mass. Representing VAT as both percentages of total %BF and absolute values (kg) allows for easier interpretation and translation to clinical relevance. Change values were calculated as the difference between follow-up and baseline measurements and were treated as continuous predictors in all models. To improve interpretability, changes in WHR were scaled by a factor of 100, so that regression coefficients in the model represent the effect of a 0.01-unit change(13,42).

#### Effect Modifiers: Baseline Adiposity

To assess whether the impact of adiposity change varied by individuals’ initial adiposity, we included corresponding baseline values (e.g., baseline weight, baseline VAT, baseline WC/WHR) in all models. These baseline values served dual roles: they were included as covariates to account for initial adiposity levels, and they were tested as effect modifiers in interaction models. Interaction terms between each change variable and its corresponding baseline measure allowed us to evaluate whether the relationship between change in adiposity and disease incidence differed by baseline adiposity level.

#### Outcome Variables: Cardiometabolic Disease CVD Biomarkers Risk Factors

Blood samples of approximately 60 mL of blood were obtained following an 8-hour fast and processed at the General Hospital Hamad Medical Centre Laboratory in Doha(25). Further details on sample collection, processing, and storage have been published(25,26). Fasting cardiometabolic biomarkers for this study included triglycerides (TG), high-density lipoprotein cholesterol (HDL-C), low-density lipoprotein cholesterol (LDL-C), total cholesterol (TC), and hemoglobin A1c (HbA1c). Duplicate systolic (SBP) and diastolic (DBP) blood pressure measurements were taken at five minute intervals with the participant in a sitting position using an Omron 705IT automated device(25). If the two measurements differed by ≥5 mm Hg a third measurement was taken, and the average of the two closest readings was used as the final SBP and DBP values in this study.

#### CVD Risk Factors

The outcomes of interest were incident cases of elevated blood pressure (EBP), type 2 diabetes, and dyslipidemia at follow-up, defined using International Diabetes Federation (IDF) guidelines and modeled as binary variables. EBP was defined as SBP of ≥130 mm Hg or DBP ≥85 mm Hg, or current use of antihypertensive medication(43). We used this threshold due to the relatively young sample and because it is part of the IDF guidelines for the definition of multiple risk factors associated with the development of cardiometabolic diseases. Diabetes was defined as HbA1c >6.5% or current use of diabetes medication, excluding participants with known type 1 diabetes (n = 16)(16). Dyslipidemia was defined as meeting one or more of the following: TC ≥5.2 mmol/L, serum TG ≥1.69 mmol/L, low HDL cholesterol (<1.03 mmol/L for males or <1.29 mmol/L for females), or current use of lipid-lowering medication(43). To focus on incident disease, participants with the respective condition at baseline were excluded from each outcome-specific analysis.

#### Covariates

All models adjusted for covariates identified a priori as potential confounders due to their known associations with both adiposity and cardiometabolic risk. These included age at baseline (continuous), duration of follow-up in years (continuous), smoking status (binary outcome of current smoker and not a smoker), and physical activity (binary outcome of physically active or not physically active). Smoking and physical activity were assessed during standardized interviews conducted by trained staff. Physical activity was measured using a shortened version of the International Physical Activity Questionnaire (IPAQ)(44) and dichotomized to distinguish between those reporting any physical activity and those reporting none, due to small cell sizes in the original categories. Similarly, smoking status was also reported as a binary variable due to small cell sizes.

#### Statistical Analysis

We compared baseline sex differences in age, smoking status, physical activity, biomarkers, anthropometric measures, body composition variables by paired t-test or McNemar’s test (**Table 1**).

**Table 1.**
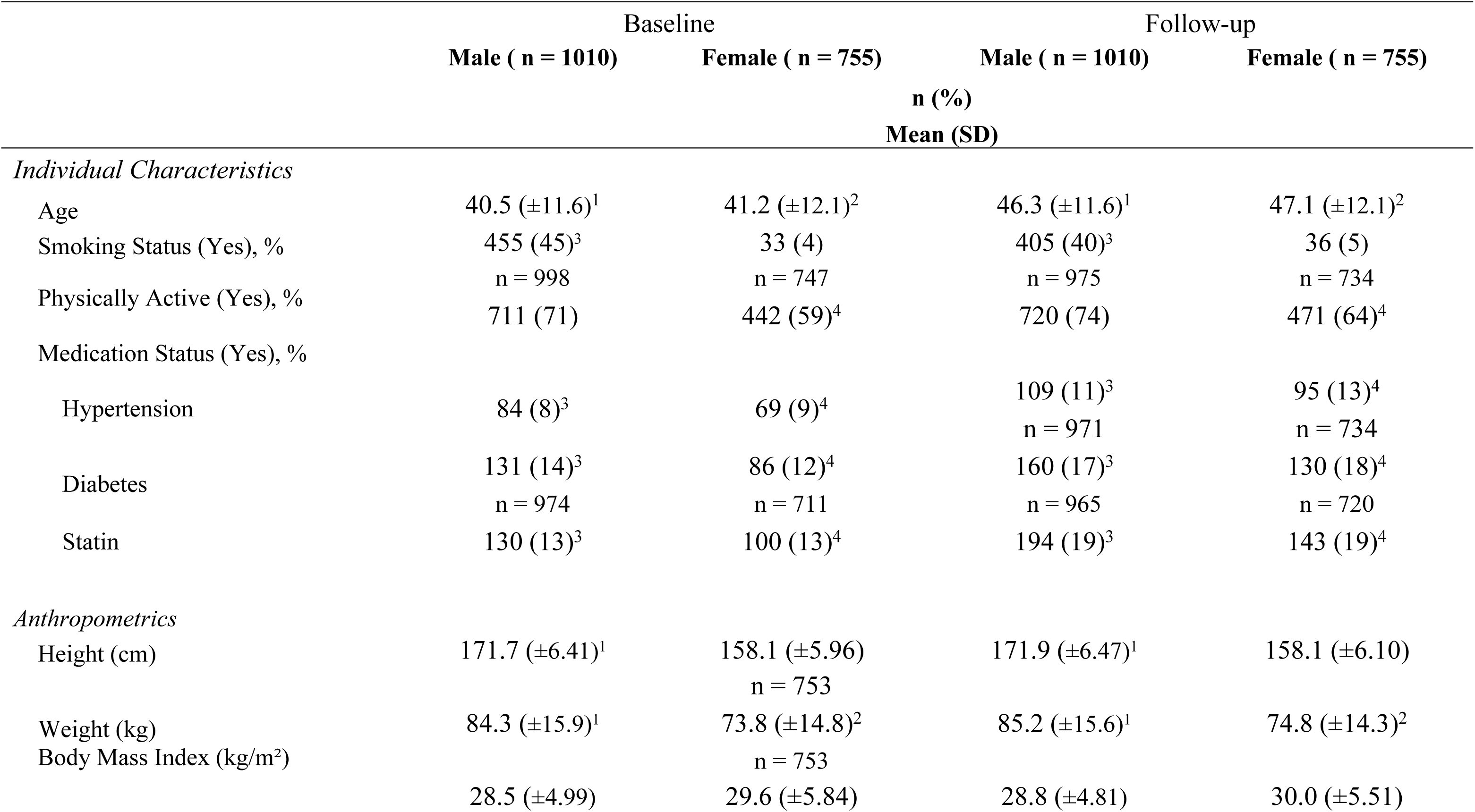
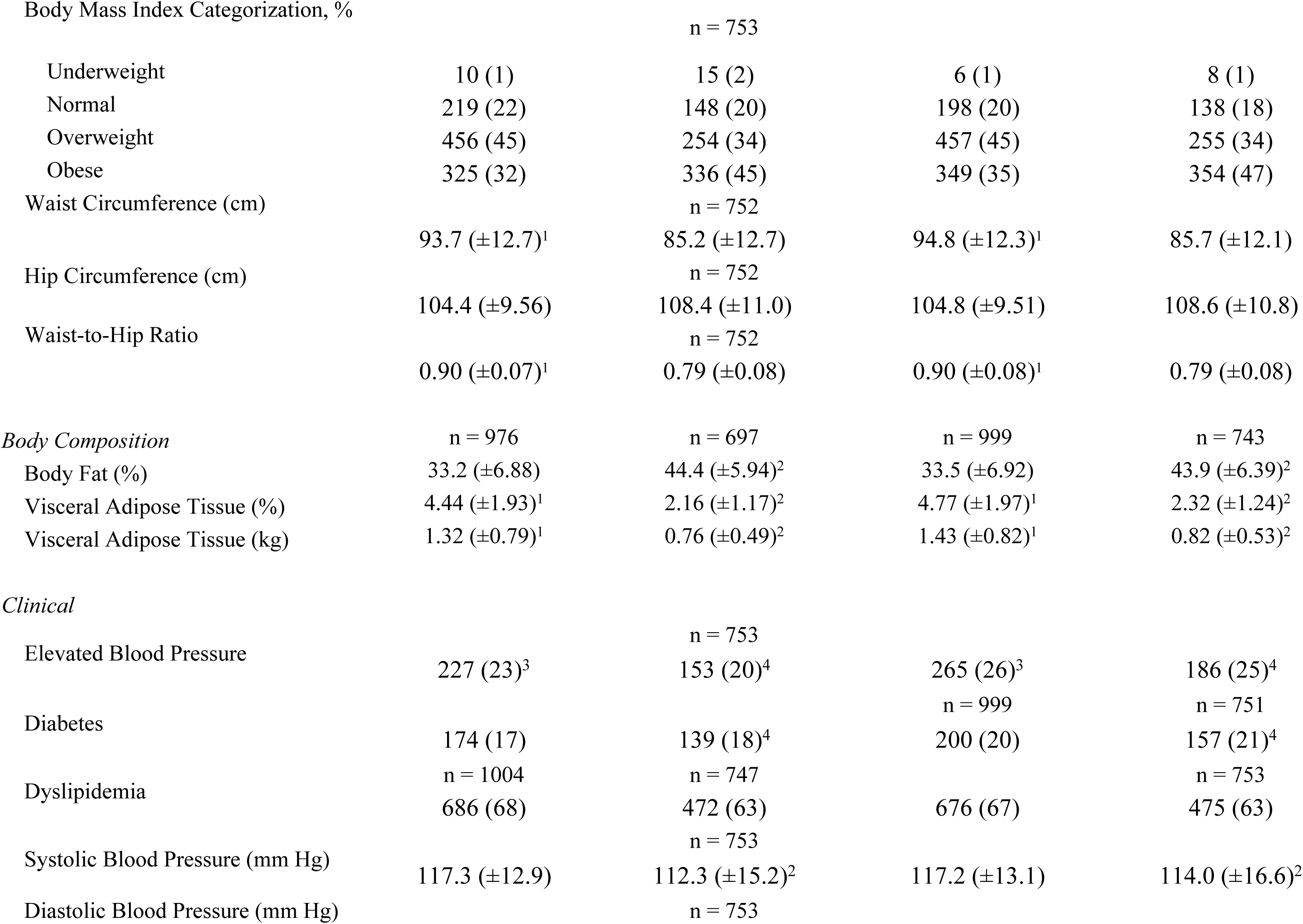
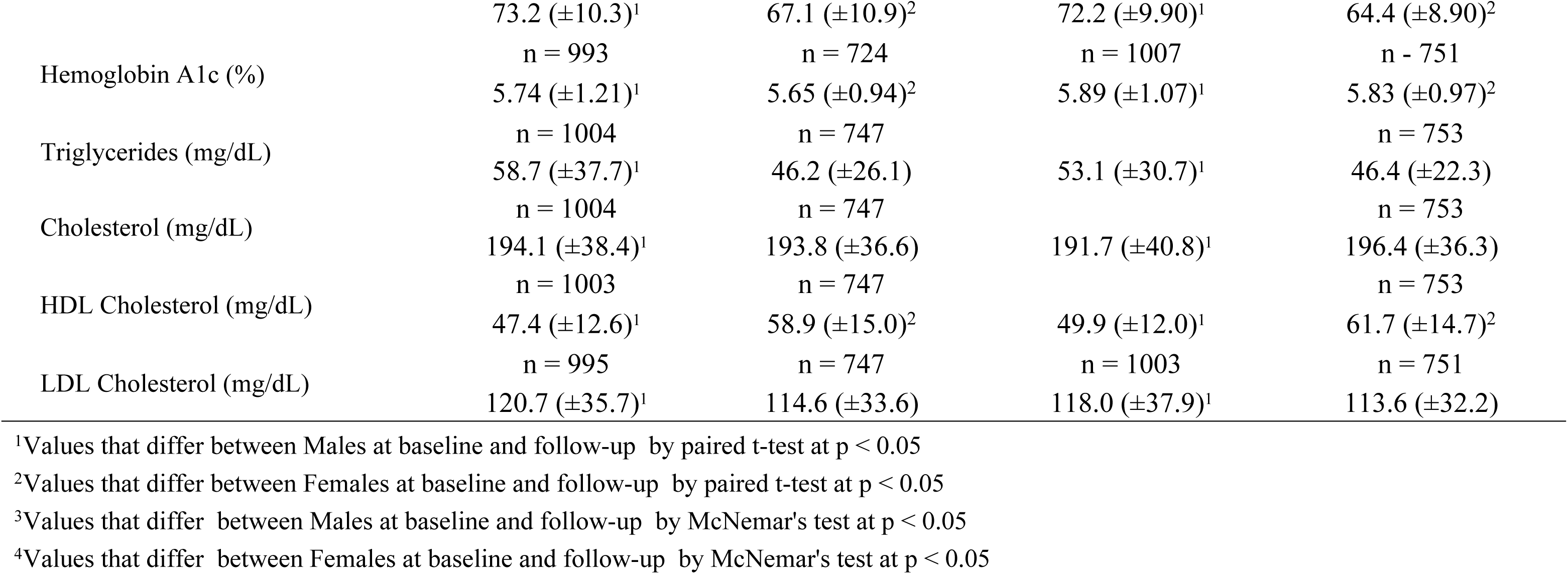
Characteristics of males and females from a Qatar BioBank cohort study sample (n = 1765).

To evaluate the relationship between changes in adiposity and incident cardiometabolic disease, we conducted sex-stratified logistic regression models for each outcome: EBP, diabetes, and dyslipidemia. Each model included a change variable (e.g., change in weight, VAT, WC, or WHR), the corresponding baseline measure, and covariates including age at baseline, duration of follow-up, smoking status, and physical activity. Additionally, we tested whether the corresponding baseline adiposity measure modified the relationship between changes in adiposity and incident disease by including interaction terms between baseline and change measures in all models. Specifically, for the change in weight model we tested whether weight gain affected incident disease risk differently according to initial weight status represented as BMI categories.

Analytic sample sizes varied across models due to differences in available follow-up data, DXA scan completeness, and disease-specific exclusions at baseline. For models assessing incident EBP, the analytic samples included 746 males and 580 females for weight and WHR/WC models, and 712 males and 525 females for VAT. For diabetes, sample sizes were 801 males and 567 females in the anthropometric models, and 768 males and 510 females for VAT. For dyslipidemia, analytic samples included 304 males and 266 females for weight and WHR/WC models, and 286 males and 241 females for VAT. All models were complete-case analyses and included only individuals free of the respective condition at baseline with valid outcome and exposure data. All analyses were conducted using STATA 18.5 (College Station, TX, USA)(45). Statistical significance was defined as *p* < 0.05. Margins and interaction plots were generated for significant interaction terms to visualize the modifying effects of baseline adiposity.

#### Sensitivity Analysis

Missing data in this study were minimal, with less than 5% missing across all variables. The sociodemographic characteristics of individuals missing anthropometric or DXA-derived data were comparable to those included in the analytic sample. Reasons for missing data included hemolyzed blood samples or inconclusive lab results (n = 19) and invalid DXA scans or participant refusal (n = 23).

Additionally, we assessed the influence of extreme values in changes of adiposity, specifically change in weight and WC. We first examined the 5th and 95th percentiles of the change variables to identify potential outliers in our longitudinal data. Extreme values were identified as likely reflecting substantial lifestyle changes, clinical interventions (e.g., bariatric surgery), longer follow-up period, or natural biological variability such as menopause-related changes. Participants with these extreme values were retained to preserve the representativeness of the sample. Sensitivity analyses were performed to assess the influence of these values.

Robust standard errors were applied to down-weight their impact, revealing no substantial differences in model results. Standardized residuals were then calculated, and observations exceeding ±3 standard deviations were excluded. Results from these sensitivity analyses were consistent with the main models, with stronger significance observed for predictors already significant. Only one additional interaction term reached significance, indicating minimal impact on our findings. Therefore, we proceeded with the standard models for interpretation.

## Results

### Participant characteristics

Among males, weight, VAT percentage and mass, and WC increased over the follow-up period. The prevalence of EBP increased from 23% at baseline to 26% at follow-up, and diabetes prevalence rose from 17% to 20% (**Table 1)**. Among females, weight, WC, and both absolute and percent VAT increased significantly over time. The prevalence of EBP increased from 20% to 25%, and diabetes from 18% to 21% **(Table 1)**. At baseline, dyslipidemia prevalence was high in both sexes, 68% in males and 63% in females, and remained similarly elevated at follow-up (67% and 63%, respectively). Incident disease cases were identified among those free of each condition at baseline. Over five years, the incidence of EBP was 4.9% in males and 5.4% in females. Diabetes incidence was 3.1% in males and 2.9% in females. Dyslipidemia incidence was modest, 1.1% in females, while prevalence slightly declined in males, suggesting possible remission or changes in clinical classification by successful treatment or lifestyle modifications, or potentially differential self-reporting.

Across all conditions, participants who developed disease were generally older at baseline than those who remained disease-free. Individuals with disease at baseline had higher BMI, WC, WHR, and VAT (**Tables 2A-C**).

**Table 2A.**
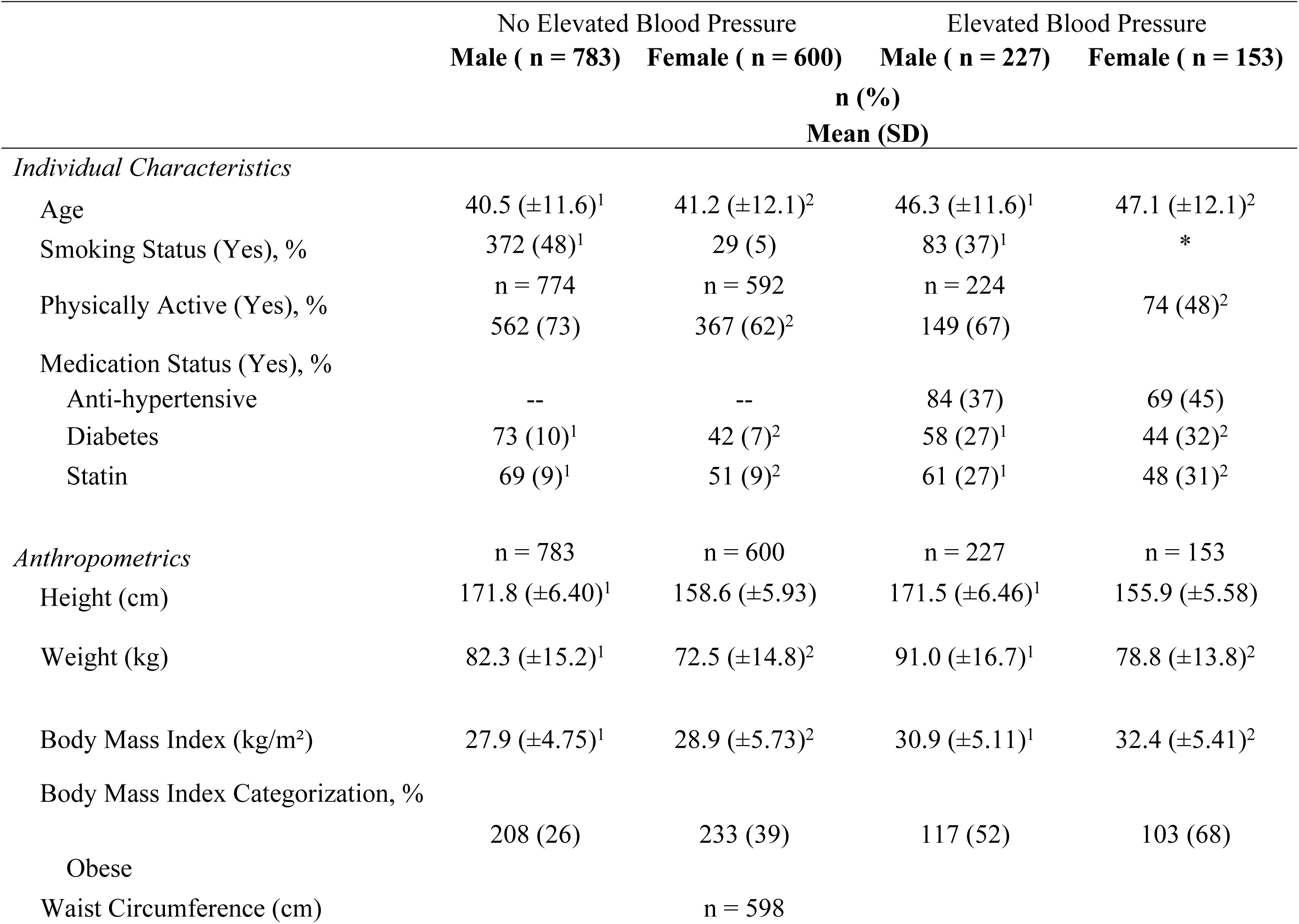
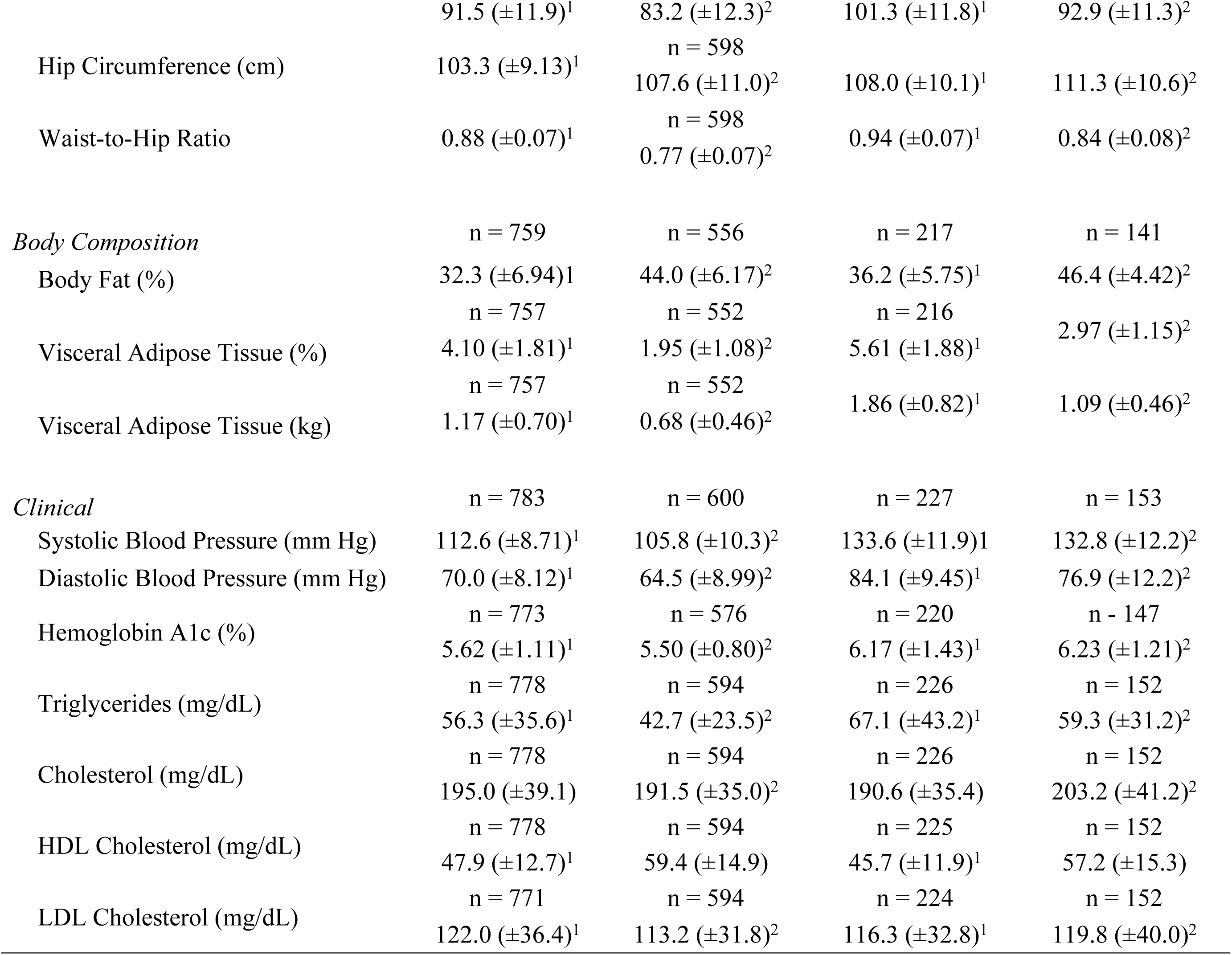
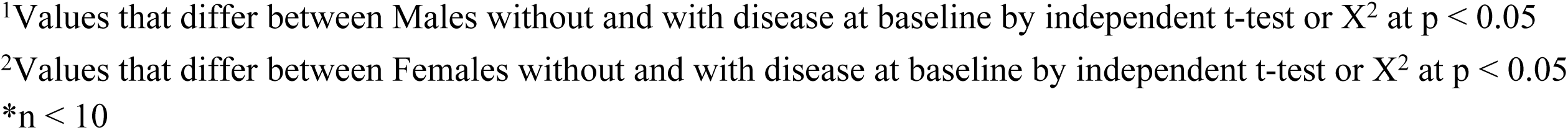
Baseline characteristics of participants with and without elevated blood pressure stratified by sex. Only individuals without the respective condition at baseline are included in the incidence analyses.

**Table 2B.**
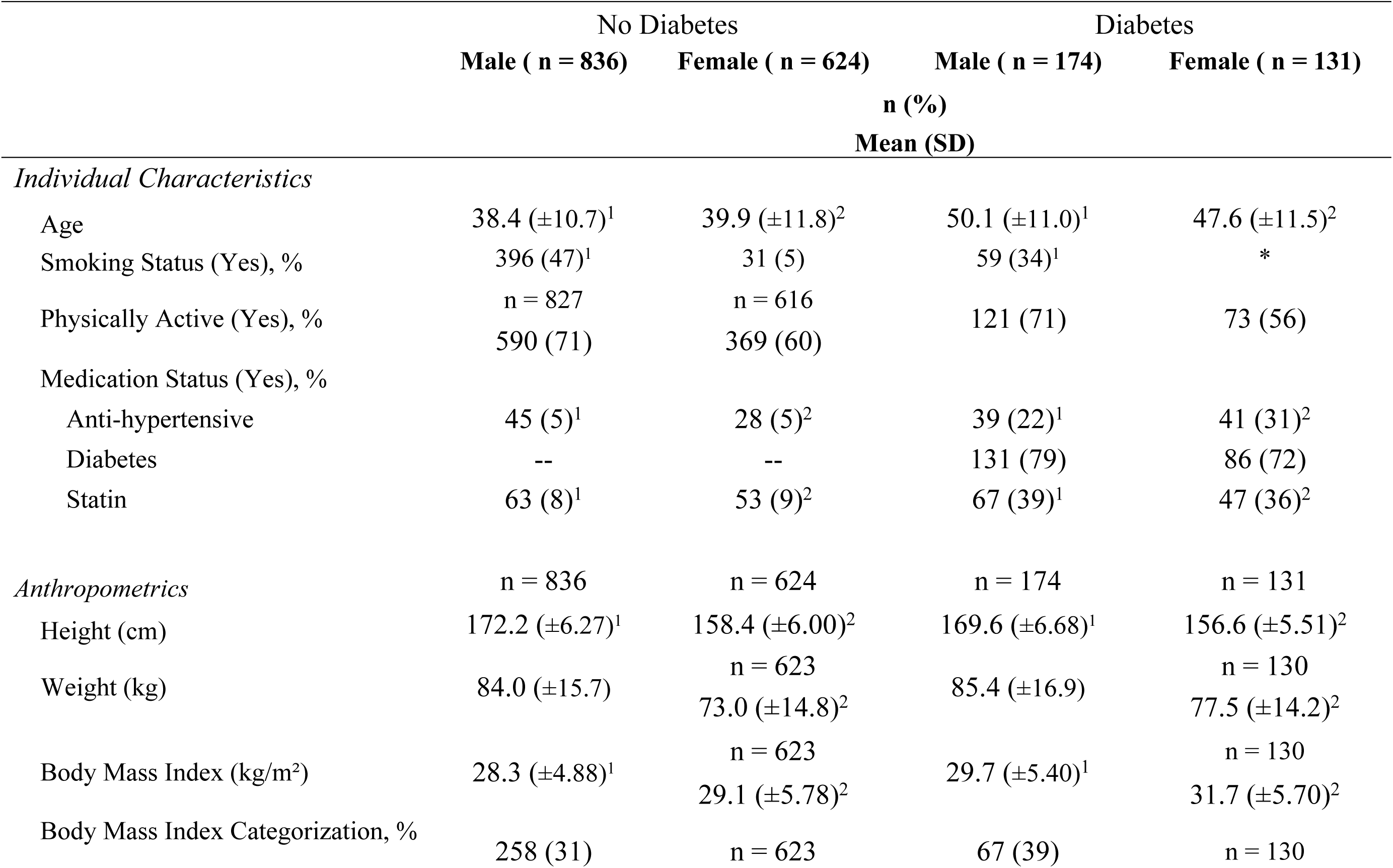
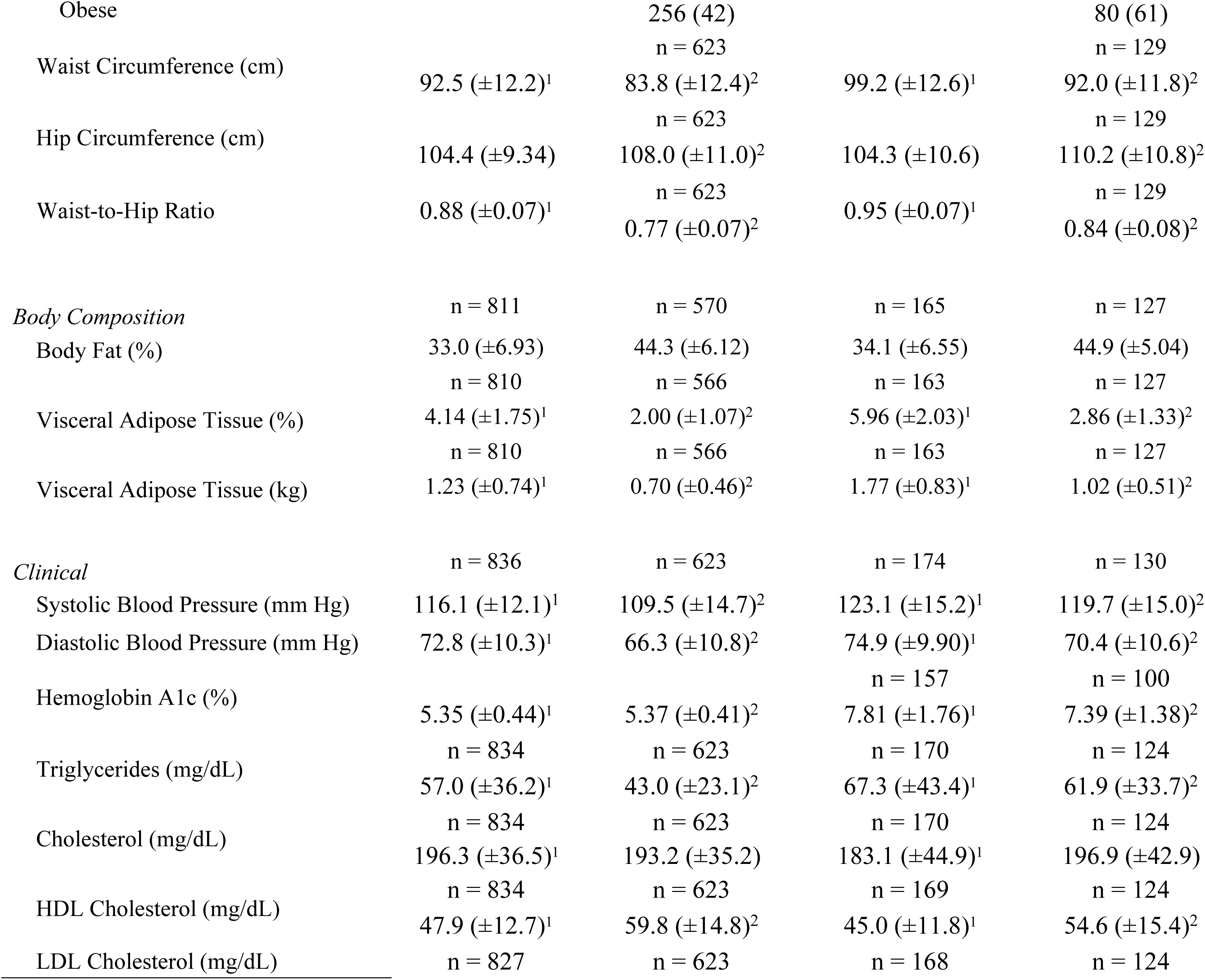
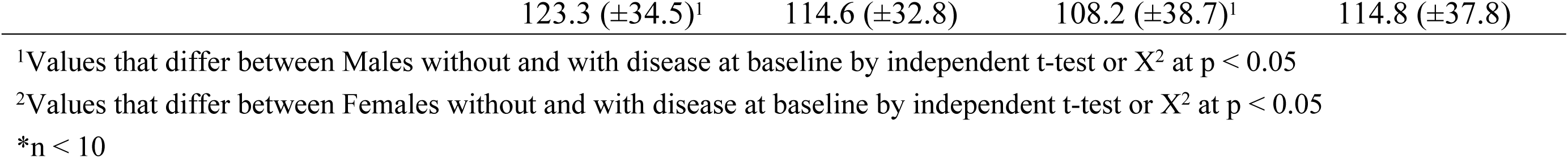
Baseline characteristics of participants with and without diabetes stratified by sex. Only individuals without the respective condition at baseline are included in the incidence analyses.

**Table 2C.**
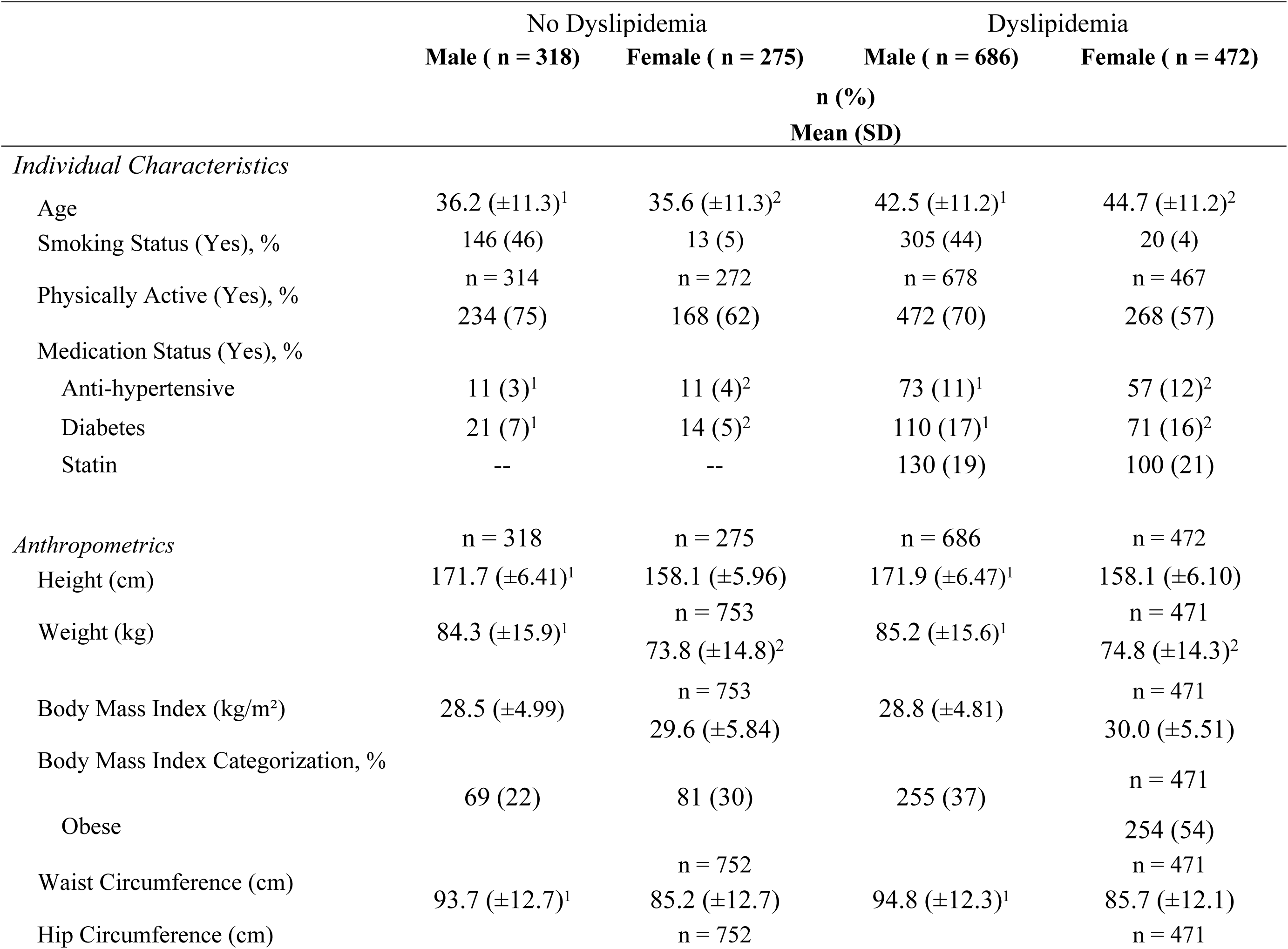
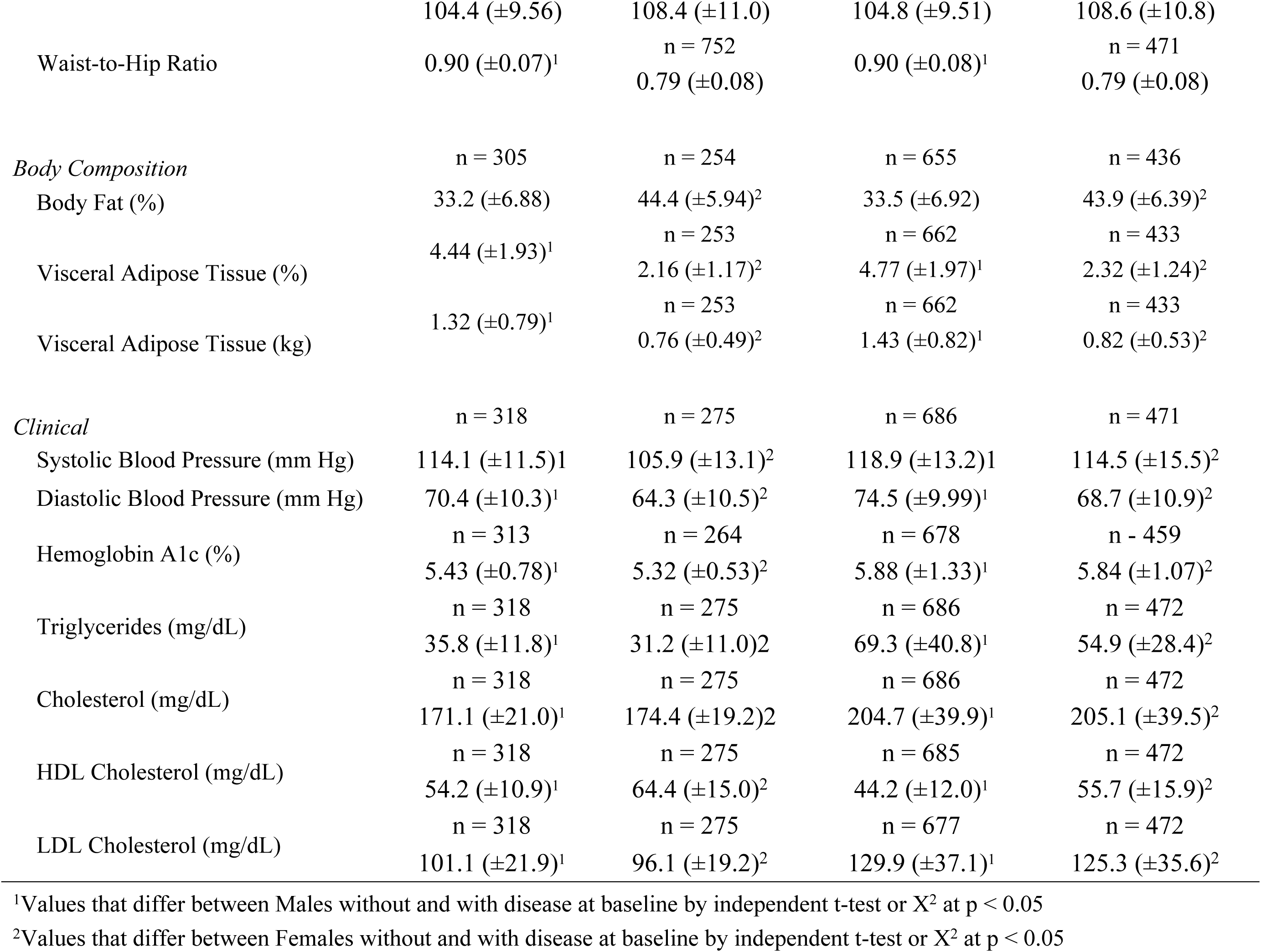
Baseline characteristics of participants with and without dyslipidemia stratified by sex. Only individuals without the respective condition at baseline are included in the incidence analyses.

|  | No Dyslipidemia |  | Dyslipidemia |  |
| --- | --- | --- | --- | --- |
|  | Male ( n = 318) | Female ( n = 275) | Male ( n = 686) | Female ( n = 472) |
|  | n (%) |  |  |  |
|  | Mean (SD) |  |  |  |
| *Individual Characteristics* |  |  |  |  |
| Age | 36.2 (±11.3)<sup>1</sup> | 35.6 (±11.3)<sup>2</sup> | 42.5 (±11.2)<sup>1</sup> | 44.7 (±11.2)<sup>2</sup> |
| Smoking Status (Yes), % | 146 (46) | 13 (5) | 305 (44) | 20 (4) |
| Physically Active (Yes), % | n = 314 | n = 272 | n = 678 | n = 467 |
|  | 234 (75) | 168 (62) | 472 (70) | 268 (57) |
| Medication Status (Yes), % |  |  |  |  |
| Anti-hypertensive | 11 (3)<sup>1</sup> | 11 (4)<sup>2</sup> | 73 (11)<sup>1</sup> | 57 (12)<sup>2</sup> |
| Diabetes | 21 (7)<sup>1</sup> | 14 (5)<sup>2</sup> | 110 (17)<sup>1</sup> | 71 (16)<sup>2</sup> |
| Statin | -- | -- | 130 (19) | 100 (21) |
| *Anthropometrics* |  |  |  |  |
|  | n = 318 | n = 275 | n = 686 | n = 472 |
| Height (cm) | 171.7 (±6.41)<sup>1</sup> | 158.1 (±5.96) | 171.9 (±6.47)<sup>1</sup> | 158.1 (±6.10) |
| Weight (kg) | 84.3 (±15.9)<sup>1</sup> | n = 753 | 85.2 (±15.6)<sup>1</sup> | n = 471 |
|  |  | 73.8 (±14.8)<sup>2</sup> |  | 74.8 (±14.3)<sup>2</sup> |
| Body Mass Index (kg/m²) | 28.5 (±4.99) | n = 753 | 28.8 (±4.81) | n = 471 |
|  |  | 29.6 (±5.84) |  | 30.0 (±5.51) |
| Body Mass Index Categorization, % |  |  |  |  |
| Obese | 69 (22) | 81 (30) | 255 (37) | n = 471 |
|  |  |  |  | 254 (54) |
| Waist Circumference (cm) | 93.7 (±12.7)<sup>1</sup> | n = 752 | 94.8 (±12.3)<sup>1</sup> | n = 471 |
|  |  | 85.2 (±12.7) |  | 85.7 (±12.1) |
| Hip Circumference (cm) |  | n = 752 |  | n = 471 |
|  | 104.4 (±9.56) | 108.4 (±11.0) | 104.8 (±9.51) | 108.6 (±10.8) |
| Waist-to-Hip Ratio | 0.90 (±0.07)<sup>1</sup> | n = 752<br>0.79 (±0.08) | 0.90 (±0.08)<sup>1</sup> | n = 471<br>0.79 (±0.08) |
| *Body Composition* | n = 305 | n = 254 | n = 655 | n = 436 |
| Body Fat (%) | 33.2 (±6.88) | 44.4 (±5.94)<sup>2</sup> | 33.5 (±6.92) | 43.9 (±6.39)<sup>2</sup> |
| Visceral Adipose Tissue (%) | 4.44 (±1.93)<sup>1</sup> | n = 253<br>2.16 (±1.17)<sup>2</sup> | n = 662<br>4.77 (±1.97)<sup>1</sup> | n = 433<br>2.32 (±1.24)<sup>2</sup> |
| Visceral Adipose Tissue (kg) | 1.32 (±0.79)<sup>1</sup> | n = 253<br>0.76 (±0.49)<sup>2</sup> | n = 662<br>1.43 (±0.82)<sup>1</sup> | n = 433<br>0.82 (±0.53)<sup>2</sup> |
| *Clinical* | n = 318 | n = 275 | n = 686 | n = 471 |
| Systolic Blood Pressure (mm Hg) | 114.1 (±11.5)<sup>1</sup> | 105.9 (±13.1)<sup>2</sup> | 118.9 (±13.2)<sup>1</sup> | 114.5 (±15.5)<sup>2</sup> |
| Diastolic Blood Pressure (mm Hg) | 70.4 (±10.3)<sup>1</sup> | 64.3 (±10.5)<sup>2</sup> | 74.5 (±9.99)<sup>1</sup> | 68.7 (±10.9)<sup>2</sup> |
| Hemoglobin A1c (%) | n = 313<br>5.43 (±0.78)<sup>1</sup> | n = 264<br>5.32 (±0.53)<sup>2</sup> | n = 678<br>5.88 (±1.33)<sup>1</sup> | n = 459<br>5.84 (±1.07)<sup>2</sup> |
| Triglycerides (mg/dL) | n = 318<br>35.8 (±11.8)<sup>1</sup> | n = 275<br>31.2 (±11.0)<sup>2</sup> | n = 686<br>69.3 (±40.8)<sup>1</sup> | n = 472<br>54.9 (±28.4)<sup>2</sup> |
| Cholesterol (mg/dL) | n = 318<br>171.1 (±21.0)<sup>1</sup> | n = 275<br>174.4 (±19.2)<sup>2</sup> | n = 686<br>204.7 (±39.9)<sup>1</sup> | n = 472<br>205.1 (±39.5)<sup>2</sup> |
| HDL Cholesterol (mg/dL) | n = 318<br>54.2 (±10.9)<sup>1</sup> | n = 275<br>64.4 (±15.0)<sup>2</sup> | n = 685<br>44.2 (±12.0)<sup>1</sup> | n = 472<br>55.7 (±15.9)<sup>2</sup> |
| LDL Cholesterol (mg/dL) | n = 318<br>101.1 (±21.9)<sup>1</sup> | n = 275<br>96.1 (±19.2)<sup>2</sup> | n = 677<br>129.9 (±37.1)<sup>1</sup> | n = 472<br>125.3 (±35.6)<sup>2</sup> |

<sup>1</sup>Values that differ between Males without and with disease at baseline by independent t-test or X<sup>2</sup> at p < 0.05
<sup>2</sup>Values that differ between Females without and with disease at baseline by independent t-test or X<sup>2</sup> at p < 0.05

### Elevated Blood Pressure (EBP)

Among males, increases in weight and WHR were significantly associated with higher odds of incident EBP (**Table 3, S1 Table**). The interaction of baseline weight with weight change was not statistically significant (**Table 4, S2 Table**). For females, weight change alone was not associated with incident EBP (**Table 3, S1 Table**); however, weight gain increased EBP risk more among those with higher baseline weight (**Table 4, S2 Table**). We used margin plots to depict this relationship which suggested that that weight loss (e.g., -10 kg) decreased EBP risk, while weight gain (+10 kg) increased risk, particularly in women with higher baseline weights

**Table 3.**
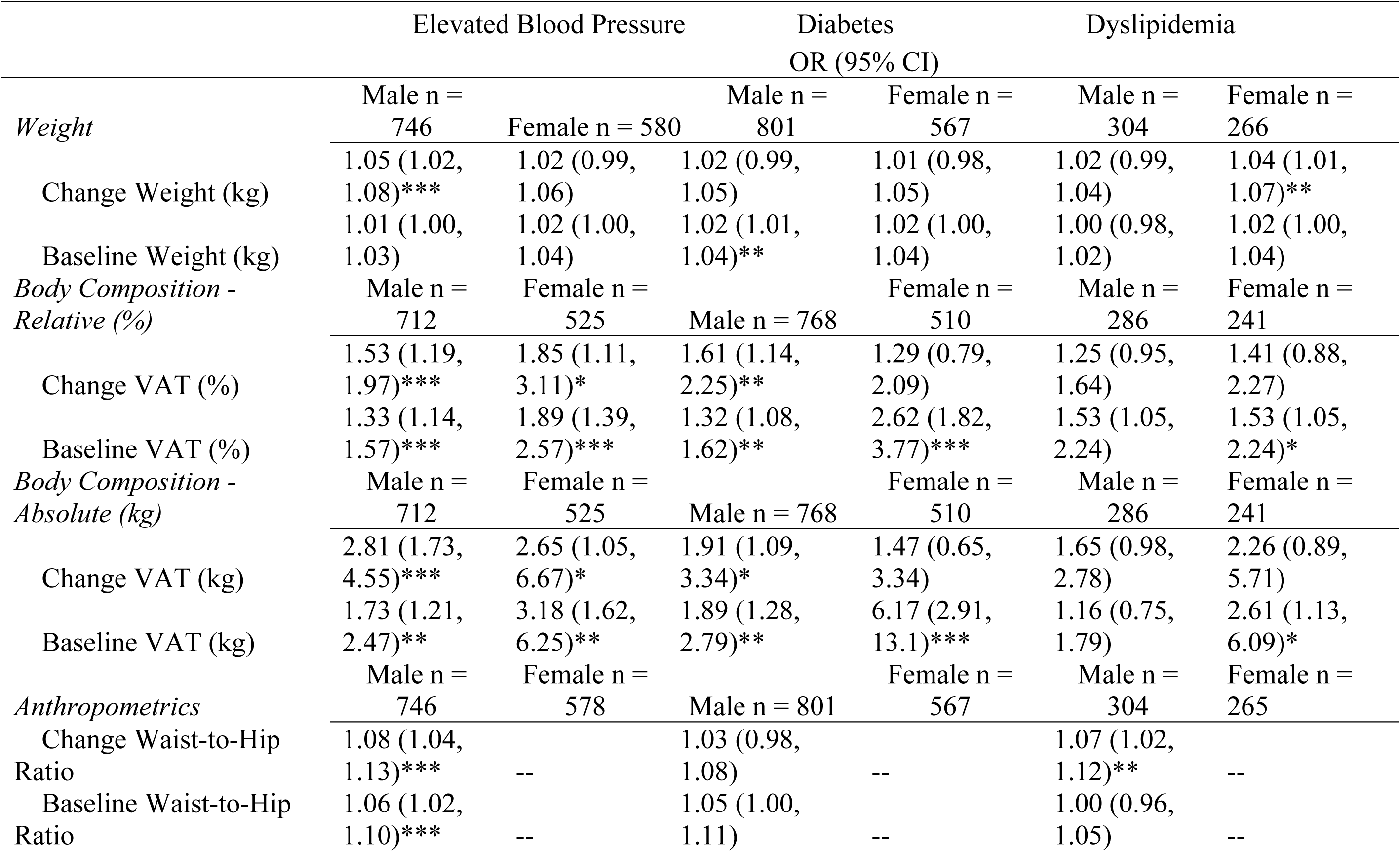
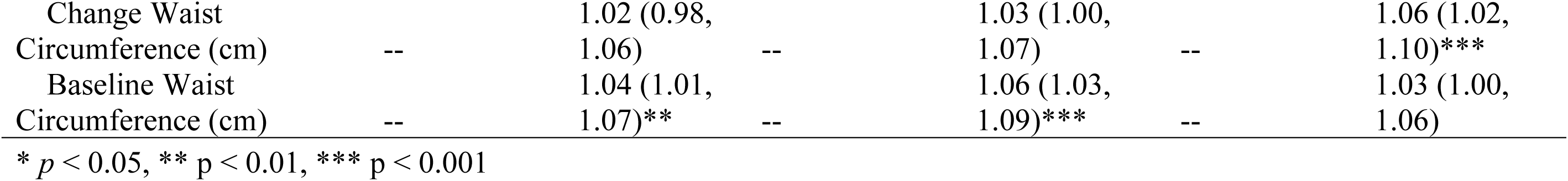
Results from incidence models assessing the relationship between changes in weight, body composition, and anthropometric measures with the incidence of elevated blood pressure, diabetes, and dyslipidemia. Odds ratios (OR) with 95% confidence intervals (CI) are presented for both male and female participants. Key variables analyzed include changes in weight, relative (%) and absolute (kg) visceral adipose tissue (VAT), waist-to-hip ratio (WHR), and waist circumference (WC), as well as their baseline measures. Models are controlled for the following covariates which include age, follow-up duration, smoking status, and physical activity. Significant associations are noted for specific changes and baseline

|  | Elevated Blood Pressure |  | Diabetes<br>OR (95% CI) |  | Dyslipidemia |  |
| --- | --- | --- | --- | --- | --- | --- |
|  | Male n =<br>746 | Female n = 580 | Male n =<br>801 | Female n =<br>567 | Male n =<br>304 | Female n =<br>266 |
| *Weight* |  |  |  |  |  |  |
| Change Weight (kg) | 1.05 (1.02, 1.08)*** | 1.02 (0.99, 1.06) | 1.02 (0.99, 1.05) | 1.01 (0.98, 1.05) | 1.02 (0.99, 1.04) | 1.04 (1.01, 1.07)** |
| Baseline Weight (kg) | 1.01 (1.00, 1.03) | 1.02 (1.00, 1.04) | 1.02 (1.01, 1.04)** | 1.02 (1.00, 1.04) | 1.00 (0.98, 1.02) | 1.02 (1.00, 1.04) |
| *Body Composition - Relative (%)* | Male n =<br>712 | Female n =<br>525 | Male n = 768 | Female n =<br>510 | Male n =<br>286 | Female n =<br>241 |
| Change VAT (%) | 1.53 (1.19, 1.97)*** | 1.85 (1.11, 3.11)* | 1.61 (1.14, 2.25)** | 1.29 (0.79, 2.09) | 1.25 (0.95, 1.64) | 1.41 (0.88, 2.27) |
| Baseline VAT (%) | 1.33 (1.14, 1.57)*** | 1.89 (1.39, 2.57)*** | 1.32 (1.08, 1.62)** | 2.62 (1.82, 3.77)*** | 1.53 (1.05, 2.24) | 1.53 (1.05, 2.24)* |
| *Body Composition - Absolute (kg)* | Male n =<br>712 | Female n =<br>525 | Male n = 768 | Female n =<br>510 | Male n =<br>286 | Female n =<br>241 |
| Change VAT (kg) | 2.81 (1.73, 4.55)*** | 2.65 (1.05, 6.67)* | 1.91 (1.09, 3.34)* | 1.47 (0.65, 3.34) | 1.65 (0.98, 2.78) | 2.26 (0.89, 5.71) |
| Baseline VAT (kg) | 1.73 (1.21, 2.47)** | 3.18 (1.62, 6.25)** | 1.89 (1.28, 2.79)** | 6.17 (2.91, 13.1)*** | 1.16 (0.75, 1.79) | 2.61 (1.13, 6.09)* |
| *Anthropometrics* | Male n =<br>746 | Female n =<br>578 | Male n = 801 | Female n =<br>567 | Male n =<br>304 | Female n =<br>265 |
| Change Waist-to-Hip Ratio | 1.08 (1.04, 1.13)*** | -- | 1.03 (0.98, 1.08) | -- | 1.07 (1.02, 1.12)** | -- |
| Baseline Waist-to-Hip Ratio | 1.06 (1.02, 1.10)*** | -- | 1.05 (1.00, 1.11) | -- | 1.00 (0.96, 1.05) | -- |
| Change Waist Circumference (cm) | -- | 1.02 (0.98, 1.06) | -- | 1.03 (1.00, 1.07) | -- | 1.06 (1.02, 1.10)*** |
| Baseline Waist Circumference (cm) | -- | 1.04 (1.01, 1.07)** | -- | 1.06 (1.03, 1.09)*** | -- | 1.03 (1.00, 1.06) |

\* $p < 0.05$ , \*\* $p < 0.01$ , \*\*\* $p < 0.001$

**Table 4.**
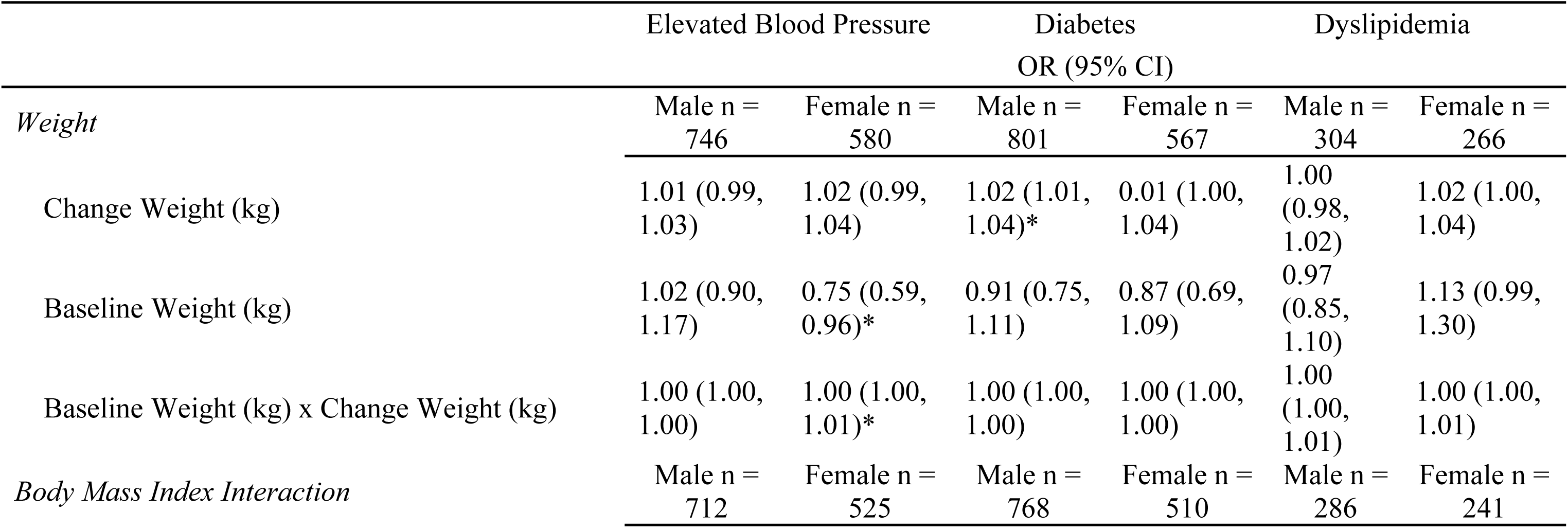
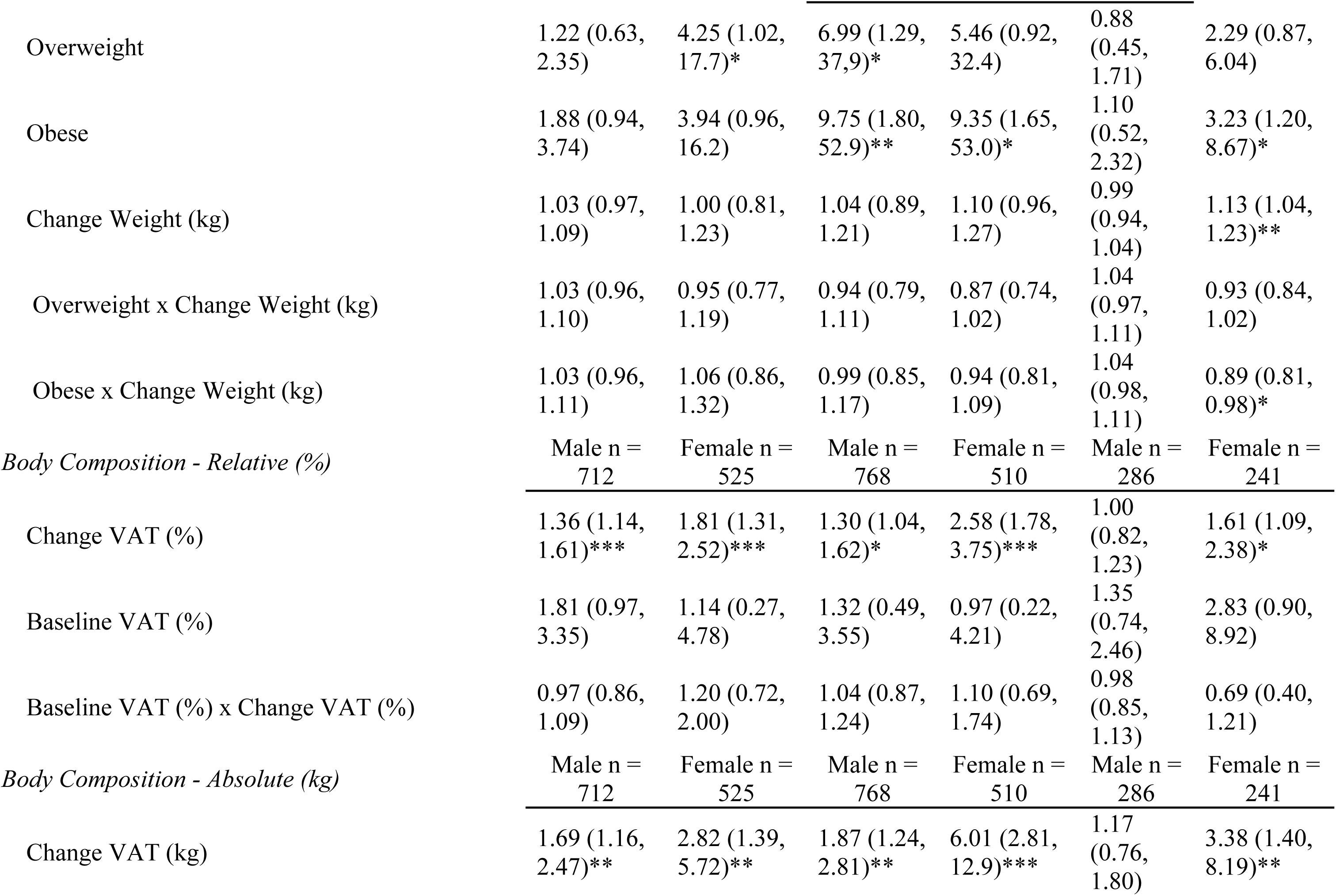
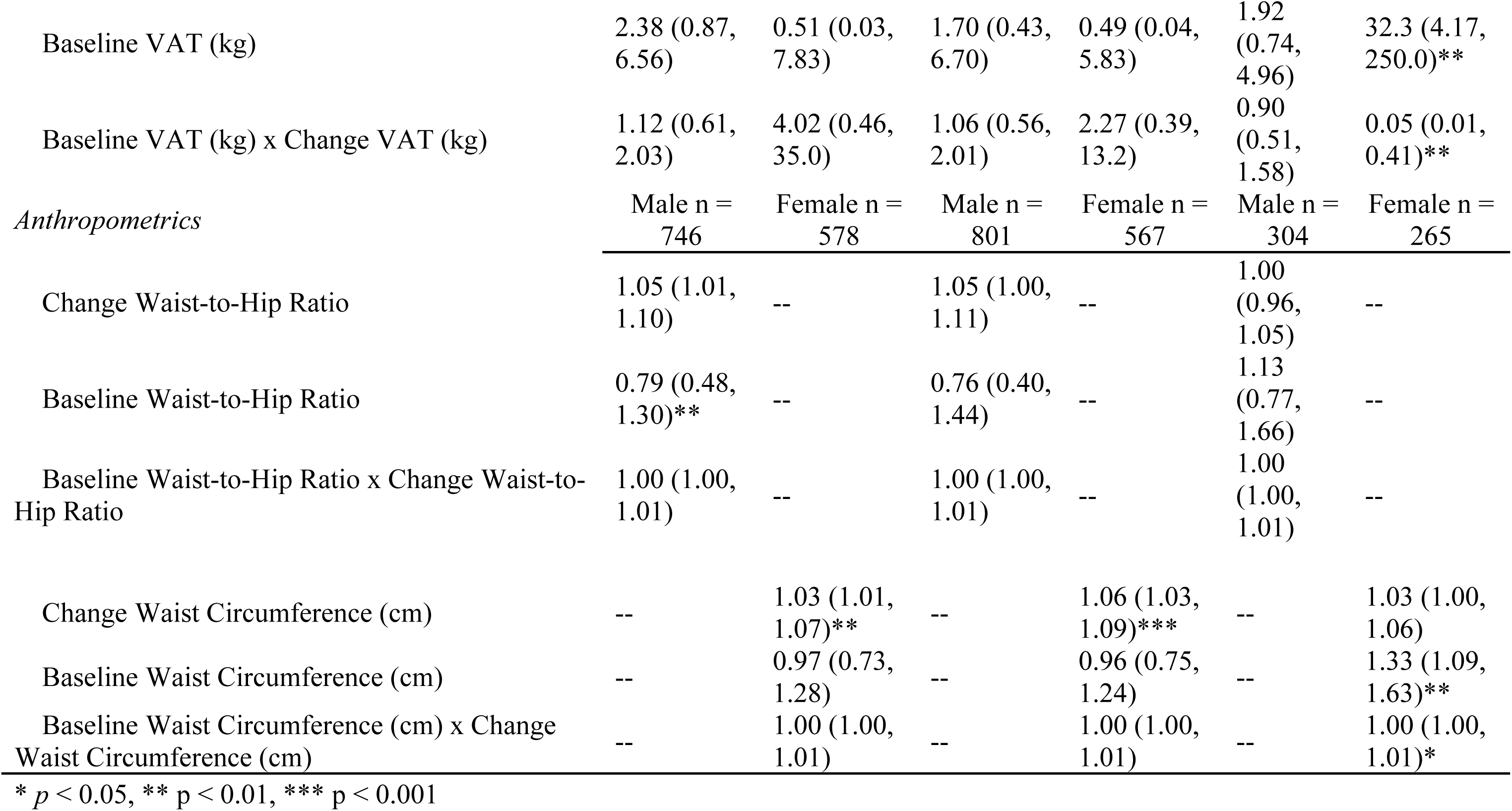
Results from interaction models assessing the combined effects of baseline and change metrics for weight, relative (%) and absolute (kg) visceral adipose tissue (VAT), WHR, WC, and BMI category (only for weight models) on the incidence of elevated blood pressure, diabetes, and dyslipidemia. Odds ratios (OR) with 95% confidence intervals (CI) are presented for both male and female participants. Models were controlled for the following covariates such as age, follow-up duration, smoking status, and physical activity. The inclusion of interaction terms provides insight into how baseline metrics moderate the relationship between changes in these variables and disease incidence. Significant interactions underscore sex-specific and metric-specific variations in disease risk.

|  | Elevated Blood Pressure |  | Diabetes<br>OR (95% CI) |  | Dyslipidemia |  |
| --- | --- | --- | --- | --- | --- | --- |
|  | Male n =<br>746 | Female n =<br>580 | Male n =<br>801 | Female n =<br>567 | Male n =<br>304 | Female n =<br>266 |
| *Weight* |  |  |  |  |  |  |
| Change Weight (kg) | 1.01 (0.99, 1.03) | 1.02 (0.99, 1.04) | 1.02 (1.01, 1.04)* | 0.01 (1.00, 1.04) | 1.00 (0.98, 1.02) | 1.02 (1.00, 1.04) |
| Baseline Weight (kg) | 1.02 (0.90, 1.17) | 0.75 (0.59, 0.96)* | 0.91 (0.75, 1.11) | 0.87 (0.69, 1.09) | 0.97 (0.85, 1.10) | 1.13 (0.99, 1.30) |
| Baseline Weight (kg) x Change Weight (kg) | 1.00 (1.00, 1.00) | 1.00 (1.00, 1.01)* | 1.00 (1.00, 1.00) | 1.00 (1.00, 1.00) | 1.00 (1.00, 1.01) | 1.00 (1.00, 1.01) |
| *Body Mass Index Interaction* | Male n =<br>712 | Female n =<br>525 | Male n =<br>768 | Female n =<br>510 | Male n =<br>286 | Female n =<br>241 |
| Overweight | 1.22 (0.63, 2.35) | 4.25 (1.02, 17.7)* | 6.99 (1.29, 37.9)* | 5.46 (0.92, 32.4) | 0.88 (0.45, 1.71) | 2.29 (0.87, 6.04) |
| Obese | 1.88 (0.94, 3.74) | 3.94 (0.96, 16.2) | 9.75 (1.80, 52.9)** | 9.35 (1.65, 53.0)* | 1.10 (0.52, 2.32) | 3.23 (1.20, 8.67)* |
| Change Weight (kg) | 1.03 (0.97, 1.09) | 1.00 (0.81, 1.23) | 1.04 (0.89, 1.21) | 1.10 (0.96, 1.27) | 0.99 (0.94, 1.04) | 1.13 (1.04, 1.23)** |
| Overweight x Change Weight (kg) | 1.03 (0.96, 1.10) | 0.95 (0.77, 1.19) | 0.94 (0.79, 1.11) | 0.87 (0.74, 1.02) | 1.04 (0.97, 1.11) | 0.93 (0.84, 1.02) |
| Obese x Change Weight (kg) | 1.03 (0.96, 1.11) | 1.06 (0.86, 1.32) | 0.99 (0.85, 1.17) | 0.94 (0.81, 1.09) | 1.04 (0.98, 1.11) | 0.89 (0.81, 0.98)* |
| *Body Composition - Relative (%)* | Male n = 712 | Female n = 525 | Male n = 768 | Female n = 510 | Male n = 286 | Female n = 241 |
| Change VAT (%) | 1.36 (1.14, 1.61)*** | 1.81 (1.31, 2.52)*** | 1.30 (1.04, 1.62)* | 2.58 (1.78, 3.75)*** | 1.00 (0.82, 1.23) | 1.61 (1.09, 2.38)* |
| Baseline VAT (%) | 1.81 (0.97, 3.35) | 1.14 (0.27, 4.78) | 1.32 (0.49, 3.55) | 0.97 (0.22, 4.21) | 1.35 (0.74, 2.46) | 2.83 (0.90, 8.92) |
| Baseline VAT (%) x Change VAT (%) | 0.97 (0.86, 1.09) | 1.20 (0.72, 2.00) | 1.04 (0.87, 1.24) | 1.10 (0.69, 1.74) | 0.98 (0.85, 1.13) | 0.69 (0.40, 1.21) |
| *Body Composition - Absolute (kg)* | Male n = 712 | Female n = 525 | Male n = 768 | Female n = 510 | Male n = 286 | Female n = 241 |
| Change VAT (kg) | 1.69 (1.16, 2.47)** | 2.82 (1.39, 5.72)** | 1.87 (1.24, 2.81)** | 6.01 (2.81, 12.9)*** | 1.17 (0.76, 1.80) | 3.38 (1.40, 8.19)** |
| Baseline VAT (kg) | 2.38 (0.87, 6.56) | 0.51 (0.03, 7.83) | 1.70 (0.43, 6.70) | 0.49 (0.04, 5.83) | 1.92 (0.74, 4.96) | 32.3 (4.17, 250.0)** |
| Baseline VAT (kg) x Change VAT (kg) | 1.12 (0.61, 2.03) | 4.02 (0.46, 35.0) | 1.06 (0.56, 2.01) | 2.27 (0.39, 13.2) | 0.90 (0.51, 1.58) | 0.05 (0.01, 0.41)** |
| *Anthropometrics* | Male n = 746 | Female n = 578 | Male n = 801 | Female n = 567 | Male n = 304 | Female n = 265 |
| Change Waist-to-Hip Ratio | 1.05 (1.01, 1.10) | -- | 1.05 (1.00, 1.11) | -- | 1.00 (0.96, 1.05) | -- |
| Baseline Waist-to-Hip Ratio | 0.79 (0.48, 1.30)** | -- | 0.76 (0.40, 1.44) | -- | 1.13 (0.77, 1.66) | -- |
| Baseline Waist-to-Hip Ratio x Change Waist-to-Hip Ratio | 1.00 (1.00, 1.01) | -- | 1.00 (1.00, 1.01) | -- | 1.00 (1.00, 1.01) | -- |
| Change Waist Circumference (cm) | -- | 1.03 (1.01, 1.07)** | -- | 1.06 (1.03, 1.09)*** | -- | 1.03 (1.00, 1.06) |
| Baseline Waist Circumference (cm) | -- | 0.97 (0.73, 1.28) | -- | 0.96 (0.75, 1.24) | -- | 1.33 (1.09, 1.63)** |
| Baseline Waist Circumference (cm) x Change Waist Circumference (cm) | -- | 1.00 (1.00, 1.01) | -- | 1.00 (1.00, 1.01) | -- | 1.00 (1.00, 1.01)* |

\* $p < 0.05$ , \*\* $p < 0.01$ , \*\*\* $p < 0.001$

In both sexes, increases in VAT percent and absolute kilograms, were significantly associated with higher odds of incident EBP (**Table 3**). No significant interactions of baseline VAT with VAT change were observed (**Table 4**). Among anthropometric predictors, increasing WHR was significantly associated with incident EBP in males, while changes in WC were not significantly associated with EBP risk in females (**Table 3**).

### Diabetes

Weight change was not significantly associated with incident diabetes in either males or females (**Table 3, S1 Table**), and no significant interactions with baseline weight were detected (**Table 4, S2 Table**). In males, increases in VAT, both relative and absolute, were significantly associated with higher odds of incident diabetes, while in females, VAT change was not a significant predictor (**Table 3**). There were no significant interactions of baseline VAT with VAT change for either sex (**Table 4**). Changes in WHR and WC were not significantly associated with incident diabetes in males or females (**Table 3 & Table 4**).

### Dyslipidemia

In females, weight gain was significantly associated with dyslipidemia incidence (**Table 5.3, S1 Table**). The interaction of BMI category with weight change was also significant (**Table 5.4, S2 Table**). The margins plot suggests that among females with obesity, additional weight gain was associated with a lower risk of dyslipidemia compared to the normal/underweight group (**Table 4, Fig 1B**). VAT change, both relative and absolute, was not significantly associated with dyslipidemia in either sex in main effects models (**Table 3**). However, in females VAT gain increased dyslipidemia risk among those with lower baseline VAT, with the effect plateauing or reversing at higher baseline levels (**Table 4, Fig 1C**). Among anthropometrics, WHR change in males and WC change in females were both significantly associated with dyslipidemia incidence (**Table 3**). In females, this association was modified by baseline WC, with dyslipidemia risk being strongest at lower baseline WC levels and diminishing as baseline WC increased (**Table 4, Figure 1D**).

**Fig 1A.**
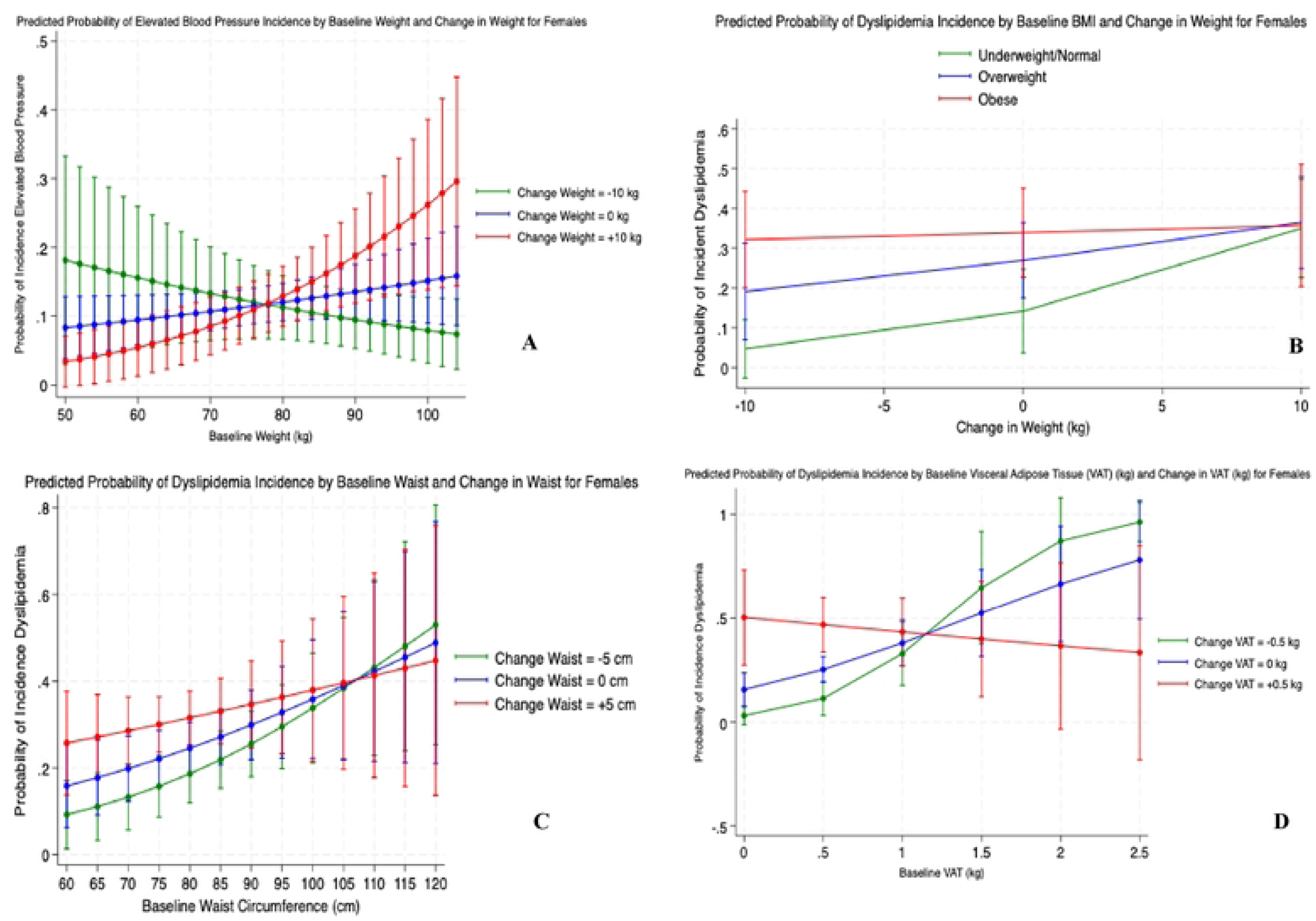
Predicted probability of disease incidence by baseline and change in adiposity measures among adults. **(A):** Elevated blood pressure (EBP) incidence by baseline weight and weight change. **(B):** Dyslipidemia incidence by body mass index (BMI) category and weight change. **(C):** Dyslipidemia incidence by baseline waist circumference and waist change. **(D):** Dyslipidemia incidence by baseline visceral adipose tissue (VAT, kg) and VAT change. All models are adjusted for age, follow-up duration, smoking status, and physical activity status. Although significant interactions were only observed in females, models were evaluated in the full sample, and results reflect significant findings stratified by sex).

## Discussion

This study examined how five-year changes in adiposity and fat distribution were associated with incident EBP, diabetes, and dyslipidemia among Qatari adults, with a focus on sex-specific associations and modification by baseline adiposity. Our findings demonstrate that changes in weight, VAT, and anthropometric proxies for VAT contribute differently to disease risk across cardiometabolic outcomes. Additionally, some of these changes and incidence of disease was modified by baseline adiposity levels.

### Participant Characteristics

While this study focused on adults with an average age of approximately 40 years, the relatively young age of the sample and the modest five-year follow-up period may have limited the observable onset of some cardiometabolic conditions. Although incidence remained low for certain outcomes, we observed increases in key clinical biomarkers such as HbA1c, total cholesterol, and blood pressure, as well as an uptick in reported use of related medications, suggesting a progression toward greater cardiometabolic risk over time (**Table 1**). The prevalence of dyslipidemia was high at baseline in both sexes, surpassing that of other outcomes and potentially indicating earlier onset of lipid-related dysfunction in this population. The elevated baseline burden of dyslipidemia may reflect unmeasured factors such as dietary patterns, genetic predispositions, or unique ethnic differences in fat metabolism that were not captured in this study. Some cardiometabolic outcomes typically emerge later in life, and as such, the age structure and follow-up duration of the sample may have contributed to the lower incidence observed and may partially explain why certain associations were not statistically significant.

### EBP

We first explored how changes in weight influence EBP risk, motivated by the well-established relationship between weight gain and hypertension(27,46–48). Our results demonstrated significant sex-specific patterns. For females, the significant interaction between baseline weight and weight change suggests that the impact of weight changes depends on initial weight status as duration of increased weight heightens the risk of chronic disease(23,24). Weight gain increased the likelihood of EBP, and weight loss was protective, especially among those at a higher weight status. Biologically, this interaction could reflect the compounding effect of adipose tissue on vascular resistance and systemic inflammation(6). Increased adiposity is associated with elevated circulating pro-inflammatory cytokines and altered vascular reactivity, contributing to hypertension(20,49). Individuals with higher baseline weight may be more sensitive to weight gain, experiencing an amplified risk of hypertension due to the additional strain placed on the cardiovascular system(23,24). This can be attributed to increased blood volume and cardiac output required to support a larger body mass, which, in turn, raises vascular resistance and blood pressure(7,8,49). Furthermore, weight gain in this group may exacerbate metabolic disturbances, such as insulin resistance and chronic inflammation, which further elevate blood pressure(7,49). While weight loss may be beneficial in this group (**Fig 1A**), further research is needed to explore or quantify the potential of weight loss, as the consequences to the cardiovascular system from prolonged exposure to excess weight might already be present. Weight loss can decrease systemic inflammation and enhance endothelial function and ultimately promote vasodilation and reducing vascular resistance(27,46). These benefits may explain the observed decrease in EBP incidence across baseline weight levels in those who lose weight. Previous studies have similarly demonstrated the link between weight changes and blood pressure, emphasizing the importance of weight management in reducing cardiometabolic risk(27,28,47,48).

Beyond weight, changes in VAT percentage and mass were significantly associated with the incidence of EBP in both sexes. These findings are consistent with the well-established role of VAT as a major driver of cardiometabolic dysfunction. VAT is metabolically active and secretes pro-inflammatory cytokines, such as interleukin-6 (IL-6) and tumor necrosis factor-alpha (TNF-α), which contribute to systemic inflammation and endothelial dysfunction(6,23,50). Differences in disease etiology likely contributed to the observed strength and consistency of VAT’s association with EBP compared to other outcomes. Hypertension is primarily influenced by increased vascular resistance, endothelial dysfunction, and arterial stiffness, all of which are exacerbated by VAT accumulation(6–8). This could explain the consistent association of VAT with EBP across sexes. Additionally, dietary factors such as high sodium intake, which were not evaluated in this study, further exacerbate fluid retention and vascular resistance(8,51). The combined effects of mechanical and hormonal mechanisms related to VAT suggest that central adiposity is a strong and biologically plausible predictor of EBP.

Among anthropometric indices, in males, WHR change was significantly associated with incident EBP, paralleling the association observed for VAT gain. WHR is thought to more precisely capture visceral fat because it attenuates the influence of subcutaneous fat by accounting for HC, which is inversely associated with cardiometabolic risk(42,52,53). In our sample, hip circumference remained relatively stable from baseline to follow-up, suggesting that changes in WHR could be driven primarily by increases in WC (**Table 1**). Prior studies have shown that both a larger waist and narrower hips are independently associated with heightened risk of hypertension, diabetes, and cardiovascular disease(52,53). WHR could be assessing the harmful effects of visceral obesity by capturing both excess abdominal fat and the loss of protective gluteofemoral mass than WC alone.

### Diabetes

In contrast to EBP, the relationship between changes in weight and incident diabetes was less consistent. Weight change was not significantly associated with incident diabetes in either sex. However, VAT gain, both relative and absolute, was significantly associated with incident diabetes in males. These findings suggest that visceral adiposity, rather than overall weight gain, may be a more relevant predictor of diabetes, particularly in men. This sex-specific pattern is consistent with existing evidence that males accumulate VAT earlier in adulthood and more extensively than females and are therefore more susceptible to VAT-driven metabolic disturbances such as insulin resistance and impaired glucose metabolism(11,20,23). More specifically, The observed association of VAT gain with diabetes in males could be influenced by biological sex differences in fat storage and metabolism(54). Hormonal differences, such as lower estrogen levels in males, might amplify the adverse effects of VAT on glucose and lipid metabolism, potentially contributing to a higher diabetes risk(8,23). Additionally, age-related declines in testosterone and loss of skeletal muscle mass have been linked to increased insulin resistance and a heightened risk of T2DM in males(55). As lean mass decreases, the relative proportion of VAT may increase, even in the absence of significant weight gain, indicating a shift toward a more metabolically detrimental body composition(55). In contrast, females are more likely to store fat in peripheral depots, such as the hips and thighs, a pattern often attributed to estrogen, which promotes gynoid fat distribution(54,56,57). This distribution pattern has been suggested to be metabolically less active than VAT and confer metabolic protection by improving blood sugar control and lipid profile, potentially buffering against the adverse effects of VAT accumulation(19,58–61). This sex-specific buffering may partly explain the lack of association between VAT and diabetes in females in this study.

Differences in disease etiology likely contributed to the observed variability in how changes in VAT influence the risk of specific diseases. Hypertension and diabetes, while both linked to metabolic dysfunction, are driven by distinct biological pathways . Diabetes is more directly driven by insulin resistance and pancreatic beta-cell dysfunction, mechanisms that are heavily influenced by ectopic fat deposition in organs like the liver and pancreas(8,51,62). The association between VAT and diabetes in males in this study may reflect their greater propensity for VAT accumulation alongside decreasing lean mass and its overall downstream metabolic effects(29,54). This hypothesis should be further tested in this sample. Furthermore, dietary factors, not assessed in this study, such as a high intake of refined carbohydrates and saturated fats further compounds insulin resistance, emphasizing the need to consider both biological and behavioral contributors to VAT-driven diabetes risk(16,17,62).

### Dyslipidemia

Dyslipidemia incidence was most strongly associated with changes in weight and anthropometric measures, such as WC, among females. In this group, weight gain significantly increased dyslipidemia risk, but this effect varied by baseline BMI. Among females with obesity, additional weight gain may not meaningfully alter risk. This pattern may reflect a saturation effect where baseline metabolic dysfunction, such as chronic inflammation and dysregulated lipid metabolism, already confers high cardiometabolic risk, and further weight gain adds little marginal risk once initial weight is taken into account(23,24).

Similarly, in females, both VAT (kg) and WC changes were significantly associated with dyslipidemia risk, but the strength of these associations varied by baseline values. VAT gain was most strongly associated with risk among women with lower baseline VAT (0 to ∼1.5 kg), consistent with VAT’s established role in metabolic dysfunction(6). However, at higher baseline VAT levels (>1.5 kg), this relationship plateaued or even reversed, with VAT loss appearing to increase dyslipidemia risk (**Fig 1D**). WC demonstrated a comparable pattern. However, sample sizes were limited at the upper extremes (∼115 cm), and confidence intervals widened substantially, indicating these findings should be interpreted cautiously (**Fig 1C**). Several biological mechanisms may explain the WC-dyslipidemia relationship. While WC is a marker of central adiposity, it does not distinguish between subcutaneous and visceral fat. Reductions in WC may reflect loss of abdominal SAT, which is relatively metabolically neutral, rather than VAT, which is more closely linked to inflammation and lipid dysregulation(30,63). If VAT remains stable or increases while SAT declines, metabolic risk may actually worsen(64,65).

Moreover, high WC is often accompanied by age-related sarcopenia and hormonal shifts in postmenopausal women, which can further contribute to dyslipidemia, even in the context of stable or decreasing waist size(9,10). Lastly, reverse causation is possible, as unintentional weight loss in individuals with advanced cardiometabolic disease may precede clinical diagnosis(66).

### Implications for Public Health

Our findings emphasize the need for targeted, multifaceted approaches to cardiometabolic disease prevention that prioritize not only weight management but also fat distribution and tailored intervention strategies. Weight management strategies should continue to prioritize individuals with higher baseline weight or those classified as overweight or obese (OW/OB). In our study, these groups demonstrated greater sensitivity to weight gain, with significant increases in disease risk, particularly for EBP among females. Conversely, weight loss in these populations, even if modest, may substantially mitigate risk. Furthermore, our findings suggested that disease risk extends beyond total weight gain to where fat is stored. As lack of availability of DXA-derived VAT estimates are a limitation in many clinical and research settings, anthropometrics that best represent VAT should be considered when screening participant or population disease risk. Incorporating these proxies into routine health assessments may enhance early detection of high-risk individuals, enabling timely, cost-effective interventions tailored to specific populations. Lastly, the suggestion of population- and sex-specific anthropometric cut-points for specific disease outcomes may offer an advantageous approach to identifying high-priority individuals for intervention. This recommendation comes from the results found in this study in which different changes in adiposity resulted in different disease incidence. By integrating these findings into practice, clinicians may be able to design interventions that account for the metabolic profiles of specific subgroups, potentially improving the effectiveness of prevention strategies.

### Challenges and Limitations

This study’s findings should be considered alongside several limitations. First, the sample consisted solely of Qatari nationals, limiting generalizability to the broader population, including expatriates. Additionally, the QBB cohort is not representative of the broader population due to its volunteer-based sampling methodology, which may introduce selection bias. Therefore, results from this study should be interpreted with caution when considering their applicability to country- or population-level health(26). Furthermore, the relatively young sample (average age 40) and short follow-up period (∼5 years) may not only limit the applicability of findings to older populations but may also help explain some of the non-significant results, given the low disease incidence for outcomes that typically develop later in life or require longer follow-up periods to detect. Small sample sizes for some covariates (smoking status, physical activity), BMI categorization of underweight participants, and outcomes necessitated the use of binary variables, reducing nuance. Definitions for EBP, diabetes, and dyslipidemia were based on the IDF criteria to align with regional studies; however, we acknowledge that alternative clinical definitions exist . Medication use was based on self-report, which, while reviewed by trained nurses at the study visit, may still be subject to misclassification. Anthropometrics were collected at a single time point per visit, and while standardized protocols were used, some degree of measurement error may remain.

### Strengths

This study had several notable strengths that enhanced the robustness and relevance of our findings. First, the use of a high-quality dataset from the QBB cohort provided comprehensive data on body composition, anthropometrics, and health metrics, enablingdetailed analyses. Second, our longitudinal design enabled the assessment of changes in weight, fat distribution, and their associations with the incidence of cardiometabolic diseases over a five-year period, providing valuable insights into dynamic risk factors rather than static measures.

Third, stratification by sex offered a nuanced understanding of how weight and fat distribution impact disease risk differently in males and females, emphasizing the importance of tailored interventions. We also incorporated anthropometric proxies like WHR and WC into our analyses, offering practical tools that can be easily applied in clinical and resource-limited settings, particularly where DXA is not feasible. Lastly, this study was conducted in a country undergoing a rapid nutrition and lifestyle transition, which may make the findings valuable for anticipating similar patterns of cardiometabolic risk in other similar countries in the region experiencing comparable shifts in diet, physical activity, and obesity prevalence.

## Conclusion

Our findings emphasize the importance of tailored approaches to cardiometabolic disease prevention that consider both weight and fat distribution. Individuals with higher baseline adiposity, particularly females, were more sensitive to weight gain, increasing their risk for conditions like EBP. Even modest weight loss in these groups may reduce risk. As DXA-based VAT estimates are often unavailable, incorporating waist-based anthropometric proxies into routine screening could improve early identification of high-risk individuals. Our findings may also support the use of population- and sex-specific cut-points, given that different adiposity changes led to varying disease risks. Integrating these insights into clinical practice may enhance the precision and effectiveness of prevention strategies. Future research should validate these results in more diverse populations and explore additional factors, such as diet and comprehensive body composition measures, to further elucidate disease risk.

## Data Availability

The data that support the findings of this study are available from the Qatar BioBank but restrictions apply to the availability of these data, which were used under license for the current study, and so are not publicly available. Data are available upon request with prior permission from the Qatar BioBank as authors do not have the permission to share the data.

## ACKNOWLEDGMENTS

Authors would like to thank Qatar University and the Grant Office for funding this work and the Qatar BioBank for providing the data used in this study. The authors are appreciative for the insightful comments by reviewers on the draft of this paper.

